# Background rates of five thrombosis with thrombocytopenia syndromes of special interest for COVID-19 vaccine safety surveillance: incidence between 2017 and 2019 and patient profiles from 25.4 million people in six European countries

**DOI:** 10.1101/2021.05.12.21257083

**Authors:** Edward Burn, Xintong Li, Kristin Kostka, Henry Morgan Stewart, Christian Reich, Sarah Seager, Talita Duarte-Salles, Sergio Fernandez-Bertolin, María Aragón, Carlen Reyes, Eugenia Martinez-Hernandez, Edelmira Marti, Antonella Delmestri, Katia Verhamme, Peter Rijnbeek, Scott Horban, Daniel R Morales, Daniel Prieto-Alhambra

## Abstract

**Background:** Thrombosis with thrombocytopenia syndrome (TTS) has been reported among individuals vaccinated with adenovirus-vectored COVID-19 vaccines. In this study we describe the background incidence of TTS in 6 European countries.

**Methods:** Electronic medical records from France, Netherlands, Italy, Germany, Spain, and the United Kingdom informed the study. Incidence rates of cerebral venous sinus thrombosis (CVST), splanchnic vein thrombosis (SVT), deep vein thrombosis (DVT), pulmonary embolism (PE), and stroke, all with concurrent thrombocytopenia, were estimated among the general population between 2017 to 2019. A range of additional adverse events of special interest for COVID-19 vaccinations were also studied in a similar manner.

**Findings:** A total of 25,432,658 individuals were included. Background rates ranged from 1.0 (0.7 to 1.4) to 8.5 (7.4 to 9.9) per 100,000 person-years for DVT with thrombocytopenia, from 0.5 (0.3 to 0.6) to 20.8 (18.9 to 22.8) for PE with thrombocytopenia, from 0.1 (0.0 to 0.1) to 2.5 (2.2 to 2.7) for SVT with thrombocytopenia, and from 0.2 (0.0 to 0.4) to 30.9 (28.6 to 33.3) for stroke with thrombocytopenia. CVST with thrombocytopenia was only identified in one database, with incidence rate of 0.1 (0.1 to 0.2) per 100,000 person-years. The incidence of TTS increased with age, with those affected typically having more comorbidities and greater medication use than the general population. TTS was also more often seen in men than women. A sizeable proportion of those affected were seen to have been taking antithrombotic and anticoagulant therapies prior to their TTS event.

**Interpretation:** Although rates vary across databases, TTS has consistently been seen to be a very rare event among the general population. While still very rare, rates of TTS are typically higher among older individuals, and those affected were also seen to generally be male and have more comorbidities and greater medication use than the general population.

**Funding:** This study was funded by the European Medicines Agency (EMA/2017/09/PE Lot 3).

## Introduction

In little over a year since the beginning of the coronavirus disease 2019 (COVID-19) pandemic, numerous vaccines against SARS-CoV-2 have been developed based on several platforms.^1^ A range of these have demonstrated a high degree of efficacy in large phase 3 clinical trials,^2–4^ received conditional approvals from regulators, and together they have already been given to over a billion individuals.^5^ The benefits of these vaccines are demonstrable with, for example, a large study on mass vaccination in Israel finding the estimated effectiveness of BNT162b2 mRNA vaccine to be 94% for symptomatic COVID-19, 87% for hospitalisation, and 92% for severe COVID-19 from seven days after the second dose.^6^ Similarly, the use of the BNT162b2 mRNA and ChAdOx1 in Scotland have been associated with substantial reductions in the risk of developing severe COVID-19 disease.^7^

There remains, however, a need to continue assessing the safety of vaccines against SARS-CoV-2 and assess safety signals as and when they arise. While the results of phase 3 clinical trials provided valuable information on the rates of relatively common, but mostly mild, adverse reactions following vaccination against SARS-CoV-2, they were not powered to study the occurrence of rare adverse events of special interest. While the risks of rare but serious adverse events might be low, nationwide vaccination campaigns where millions of people are inoculated can lead to a considerable absolute number of any such events to occur.

A particular area of concern has recently arisen relating to the occurrence of thrombosis (often cerebral or abdominal) with concomitant thrombocytopenia among individuals who had received adenovirus-based vaccine against SARS-CoV-2. As of the 28^th^ April 2021, 242 instances of thromboembolic events with thrombocytopenia among individuals who had recently received the ChAdOx1 vaccine in the UK had been identified on the basis of spontaneous reports. Of these, cerebral venous sinus thrombosis (CVST) was reported in 93 of the cases.^8^ Meanwhile, as of the 23^rd^ April 2021, 15 confirmed reports of thrombosis with thrombocytopenia syndrome (TTS) had been identified for the Ad.26.COV2.S vaccine in the US.^9^ These spontaneous reports of TTS among individuals who had received the ChAdOx1 and Ad.26.COV2.S vaccines came at a time when 22.6 million first doses and 5.9 million second doses of the ChAdOx1 vaccine had been given in the UK and more than 8 million doses of the Ad.26.COV2.S had been given in the US.^8,9^

The degree to which the reported TTS events exceed the number that would typically be observed in the absence of vaccinations against SARS-CoV-2 is not yet well-known, nor is how the profiles of those with such events after vaccination have differed from those who typically experience them. Establishing the rates of TTS events among the general population in previous years will help to provide context for the observed rates being seen among those vaccinated.^10^ Moreover, a description of the profiles of the individuals who have had TTS events in the past will also help to inform a consideration of whether the profiles of individuals with TTS after a vaccination against COVID-19 differ to those who typically have such events.

In this study we set out to estimate the background incidence rates of TTS and to describe the profiles of individuals who typically have these types of events. We did this using electronic medical records collected between 2017 to 2019 and covering over 25 million people across six European countries. In addition, we performed similar analyses for a range of other embolic and thrombotic events and coagulopathies of special interest for COVID-19 vaccinations.

## Methods

### Study Design

An international network cohort study using routinely-collected primary care data from across Europe. Data were previously mapped to the Observational Medical Outcomes Partnership (OMOP) Common Data Model (CDM), which allowed for the study to be run in a distributed manner, with common analytic code run by each site without the need to share patient-level data between sites.^11–13^

### Data sources

Data from seven electronic medical records databases from France, Netherlands, Italy, Germany, Spain, and the United Kingdom informed the analysis. The Clinical Practice Research Datalink (CPRD) GOLD database contains data contributed by general practitioners (GP) from the United Kingdom.^14^ The Health Informatics Centre at the University of Dundee (HIC Dundee) database includes linked primary care and hospital data for persons from the Tayside region of Scotland, capturing around 20% of the Scottish population. The Integrated Primary Care Information (IPCI) database is collected from electronic healthcare records of patients registered with GPs throughout the Netherlands. IQVIA Longitudinal Patient Data (LPD) Italy includes anonymised patient records collected from software used by GPs during an office visit to document patients’
s clinical records. IQVIA LPD France is a computerised network of physicians including GPs who contribute to a centralised database of anonymised patient electronic medical records.^15^ IQVIA Disease Analyser (DA) Germany is collected from extracts of patient management software used by GPs and specialists practicing in ambulatory care settings. The Information System for Research in Primary Care (SIDIAP; www.sidiap.org) is a primary care records database that covers approximately 80% of the population of Catalonia, North-East Spain. SIDIAP can be linked to the minimum basic set of hospital discharge data (CMBD-HA), which includes diagnosis and procedures registered during hospital admissions.^16^ Results for SIDIAP CMBD-HA are presented in this manuscript, with results for SIDIAP only also reported in the supplementary materials for comparison.

In summary, all the included databases captured outpatient diagnoses and lab measurements. SIDIAP CMBDH-HA and HIC Dundee also directly captured diagnoses from linked hospital data. HIC Dundee was the only database that, in addition, included hospital lab measurements. Study participants and time at risk

The primary study cohort consisted of individuals present in a database as of the 1^st^ January 2017, with this date used as the index date for all study participants. These individuals were followed up to whichever came first: the outcome of interest, exit from the database, or the 31^st^ December 2019 (the end of study period). A second study cohort definition of active patients was considered, where individuals entered the cohort on the date of their first visit occurrence after 1st January 2017. As with the primary study cohorts these individuals were followed up to whichever came first: the outcome of interest, exit from the database, or 31^st^ December 2019. As a sensitivity analysis, study cohorts were also generated with the additional requirement that individuals had a minimum of one year of history available in the database prior to their index date. For each analysis of a given outcome, any individual with that outcome in the year prior to their index date was excluded.

### Outcomes

Here we summarise results for five specific TTS events of interest: CVST with thrombocytopenia, DVT with thrombocytopenia, PE with thrombocytopenia, SVT with thrombocytopenia, and stroke with thrombocytopenia. Occurrences of CVST, DVT, PE, and stroke were identified on the basis of diagnostic codes. Given the limited granularity in coding for a number of the databases, stroke encapsulated both ischemic and hemorrhagic stroke. Thrombocytopenia was identified either by a diagnostic code or a measurement of between 10,000 to 150,000 platelets per microliter of blood and was required to have been observed over a time window starting ten days prior to the event of interest up to ten days afterwards. For comparison, we also provide results for each of the outcomes without the need for them to be accompanied by thrombocytopenia. In addition, we provide background rates for coagulopathies that have been identified as potential causes for TTS: heparin-induced thrombocytopenia (HIT), disseminated intravascular coagulation (DIC), immune thrombocytopenia, and thrombotic thrombocytopenic purpura (TTP - which included hemolytic uremic syndrome).

The outcomes described here are taken from a wider set of adverse events of special interest (AESI) for COVID-19 vaccinations. Three sets of outcome events were identified: 1) venous thromboembolic events, 2) arterial thromboembolic events, and 3) rare embolic, coagulopathies, and TTS events. For venous thromboembolic events, instances of DVT (with one broad definition and another narrow - in this manuscript, we describe results for the narrow definition) and PE events were identified, with venous thromboembolism events defined as the occurrence of either DVT or PE. For arterial thromboembolic events, instances of myocardial infarction and ischemic stroke were identified, along with the composite outcome of either of these events. Instances of stroke, either ischemic or hemorrhagic, were also identified. A wide set of rare embolic and thrombotic events and thrombocytopenias and platelet disorders were considered: DIC, immune thrombocytopenia, TTP, HIT, thrombocytopenia, thrombotic thrombocytopenic purpura, platelet disorder/s, CVST, splenic vein thrombosis, splenic artery thrombosis, splenic infarction, hepatic vein thrombosis, hepatic artery thrombosis, portal vein thrombosis, intestinal infarction, mesenteric vein thrombosis, celiac artery thrombosis, visceral vein thrombosis, and SVT.

All study outcomes were identified using code lists reviewed by three epidemiologists (EB, DPA, CR), a clinical neurologist (EMH) and a clinical haematologist (EM). These definitions were reviewed using the aid of the CohortDiagnostics R package,^17^ so as to identify additional codes of interest and to remove those highlighted as irrelevant based on feedback from regulators (e.g. puerperium and pregnancy-related disease) through an iterative process during the initial stages of analyses. A detailed description of the definitions used to identify the outcomes of the study is provided at https://livedataoxford.shinyapps.io/CovCoagOutcomesCohorts/. This application summarises the codes used to identify outcomes and their frequency in the databases used in the study, the overlap between cohorts in the databases as a whole, and a detailed summary of the profiles of all the individuals with a code of interest in each of the databases.

### Patient profiles

The characteristics of the study population were extracted relative to their index date, as were those of individuals with a particular outcome of interest relative to the date of their event. The age and sex of individuals was identified, along with their history of conditions and medication use. Using all of an individual’s prior observation time, prior diagnosis of autoimmune disease, antiphospholipid syndrome, thrombophilia, asthma, atrial fibrillation, malignant neoplastic disease, diabetes mellitus, obesity, heart disease, hypertensive disorder, renal impairment, chronic obstructive pulmonary disease (COPD), or dementia were identified. Meanwhile, prior medication use was identified using a time window of 183 days prior to four days prior the index date. Drug exposures of antithrombotic and anticoagulant therapies, non-steroidal anti-inflammatory drugs, Cox-2 inhibitors, systemic corticosteroids, lipid modifying agents, antineoplastic and immunomodulating agents, hormonal contraceptives for systemic use, tamoxifen, and sex hormones and modulators of the genital system were identified over this time window.

### Statistical methods

The profiles of the study cohorts and those with an outcome of interest were summarised, with median and interquartile range (IQR) used for continuous variables and counts and percentages used for categorical variables. For each study outcome, the number of events, the observed time at risk, and the incidence rate per 100,000 person-years are summarised along with 95% confidence intervals. These results are provided for the study cohorts and stratified by data source as a whole and stratified by age (≤44, 45 to 64, or ≥65 years old) and sex. To aid in comparison with rates being reported after vaccinations, the expected number of events per 28 days for a population of 10 million were calculated based on the incidence rates calculated for the overall study cohorts and age strata.

### Code availability

All analytic code used for the analysis has been made publicly available at https://github.com/oxford-pharmacoepi/CovCoagBackgroundIncidence.

## Results

A total of 25,432,658individuals were included in the study (3,913,071 from CPRD, 948,561 from HIC Dundee, 8,459,098 DA Germany, 3,951,633 LPD France, 1,299,288 IPCI, 1,066,230 LPD Italy, and 5,794,777 from SIDIAP CMBDH-HA). The median age of the background populations ranged from 41 in CPRD to 52 in DA Germany and LPD Italy. More detailed characteristics of each of these populations are summarised in Table 1.

**Table 1:** Characteristics of study populations

|  | CPRD | HIC Dundee | IPCI<br>The<br>Netherlands | IQVIA DA<br>Germany | IQVIA LPD<br>France | IQVIA LPD<br>Italy | SIDIAP<br>CMBDH-HA |
| --- | --- | --- | --- | --- | --- | --- | --- |
| N | 3,913,071 | 948,561 | 1,299,288 | 8,459,098 | 3,951,633 | 1,066,230 | 5,794,777 |
| Age (Median [IQR]) | 41 [22 to 59] | 41 [23 to 59] | 44 [23 to 60] | 52 [32 to 67] | 48 [28 to 65] | 52 [37 to 68] | 42 [25 to 59] |
| Sex: Male (N [%]) | 1,937,858<br>(49.5%) | 469,725<br>(49.5%) | 636,386<br>(49.0%) | 3,589,506<br>(42.4%) | 1,669,415<br>(42.2%) | 426,758<br>(40.0%) | 2,859,044<br>(49.3%) |
| Years of prior observation time (Median [IQR]) | 11.9 [4.7 to 15.1] | 8.0 [6.6 to 12.0] | 3.2 [1.8 to 5.7] | 4.8 [1.9 to 8.9] | 4.6 [2.0 to 6.2] | 6.3 [5.0 to 6.5] | 11.0 [11.0 to 11.0] |
| Comorbidities |  |  |  |  |  |  |  |
| Autoimmune disease (N [%]) | 70,604 (1.8%) | 8,040 (0.8%) | 24,645 (1.9%) | 238,985<br>(2.8%) | 32,245 (0.8%) | 45,567<br>(4.3%) | 84,817 (1.5%) |
| Antiphospholipid syndrome (N [%]) | 1,166 (0.0%) | <5 | <5 | <5 | <5 | <5 | 1,011 (0.0%) |
| Thrombophilia (N [%]) | 3,039 (0.1%) | 198 (0.0%) | 0 (0.0%) | 6,474 (0.1%) | 313 (0.0%) | <5 | 2,796 (0.0%) |
| Asthma (N [%]) | 484,991<br>(12.4%) | 37,160 (3.9%) | 138,777<br>(10.7%) | 412,789<br>(4.9%) | 222,161<br>(5.6%) | 79,528<br>(7.5%) | 353,485<br>(6.1%) |
| COPD (N [%]) | 80,393 (2.1%) | 14,225 (1.5%) | 40,116 (3.1%) | 358,047<br>(4.2%) | 41,040 (1.0%) | 27,119<br>(2.5%) | 166,817<br>(2.9%) |
| Atrial fibrillation (N [%]) | 76,091 (1.9%) | 825 (0.1%) | 31,801 (2.4%) | 92,767 (1.1%) | 13,412 (0.3%) | 34,325<br>(3.2%) | 137,843<br>(2.4%) |
| Diabetes mellitus (N [%]) | 213,996<br>(5.5%) | 25,891 (2.7%) | 93,035 (7.2%) | 597,233<br>(7.1%) | 174,564<br>(4.4%) | 95,611<br>(9.0%) | 468,808<br>(8.1%) |
| Obesity (N [%]) | 107,522<br>(2.7%) | 9,843 (1.0%) | 40,395 (3.1%) | 530,958<br>(6.3%) | 15,634 (0.4%) | 46,101<br>(4.3%) | 927,483<br>(16.0%) |
| Heart disease (N [%]) | 278,323<br>(7.1%) | 56,966 (6.0%) | 129,562<br>(10.0%) | 936,730<br>(11.1%) | 194,630<br>(4.9%) | 165,172<br>(15.5%) | 592,122<br>(10.2%) |
| Hypertensive disorder (N [%]) | 558,671<br>(14.3%) | 71,703 (7.6%) | 222,433<br>(17.1%) | 1,425,782<br>(16.9%) | 498,244<br>(12.6%) | 322,776<br>(30.3%) | 1,145,518<br>(19.8%) |
| Renal impairment (N [%]) | 168,610<br>(4.3%) | 17,311 (1.8%) | 27,555 (2.1%) | 169,166<br>(2.0%) | 13,064 (0.3%) | 31,853<br>(3.0%) | 230,896<br>(4.0%) |
| Malignant neoplastic disease (N [%]) | 198,275<br>(5.1%) | 51,307 (5.4%) | 106,223<br>(8.2%) | 534,352<br>(6.3%) | 66,962 (1.7%) | 86,645<br>(8.1%) | 342,511<br>(5.9%) |
| Dementia (N [%]) | 33,537 (0.9%) | 5,515 (0.6%) | 7,873 (0.6%) | 95,957 (1.1%) | 9,217 (0.2%) | 10,458 (1.0%) | 72,696 (1.3%) |
| Medication use (183 days prior to four days prior) |  |  |  |  |  |  |  |
| Non-steroidal anti-inflammatory drugs (N [%]) | 900,092 (23.0%) | 247,182 (26.1%) | 211,464 (16.3%) | 928,497 (11.0%) | 1,056,021 (26.7%) | 293,188 (27.5%) | 1,617,103 (27.9%) |
| Cox-2 inhibitors (N [%]) | 7,126 (0.2%) | 2,469 (0.3%) | 8,165 (0.6%) | 33,006 (0.4%) | 13,769 (0.3%) | 21,899 (2.1%) | 27,048 (0.5%) |
| Systemic corticosteroids (N [%]) | 404,443 (10.3%) | 95,032 (10.0%) | 139,482 (10.7%) | 269,020 (3.2%) | 315,054 (8.0%) | 84,587 (7.9%) | 337,121 (5.8%) |
| Antithrombotic and anticoagulant therapies (N [%]) | 114,246 (2.9%) | 72,557 (7.6%) | 44,985 (3.5%) | 213,378 (2.5%) | 175,535 (4.4%) | 110,079 (10.3%) | 112,901 (1.9%) |
| Lipid modifying agents (N [%]) | 143,424 (3.7%) | 103,695 (10.9%) | 54,039 (4.2%) | 191,407 (2.3%) | 197,031 (5.0%) | 83,234 (7.8%) | 81,743 (1.4%) |
| Antineoplastic and immunomodulating agents (N [%]) | 124,080 (3.2%) | 44,853 (4.7%) | 54,941 (4.2%) | 210,390 (2.5%) | 94,702 (2.4%) | 36,792 (3.5%) | 64,163 (1.1%) |
| Hormonal contraceptives for systemic use (N [%]) | 173,708 (4.4%) | 51,084 (5.4%) | 47,983 (3.7%) | 169,549 (2.0%) | 98,852 (2.5%) | 18,740 (1.8%) | 46,834 (0.8%) |
| Tamoxifen (N [%]) | 2,141 (0.1%) | 1,666 (0.2%) | 865 (0.1%) | 3,761 (0.0%) | 826 (0.0%) | 684 (0.1%) | 1,230 (0.0%) |
| Sex hormones and modulators of the genital system (N [%]) | 213,023 (5.4%) | 63,019 (6.6%) | 55,810 (4.3%) | 228,846 (2.7%) | 141,501 (3.6%) | 29,750 (2.8%) | 58,987 (1.0%) |

CPRD: Clinical Practice Research Datalink, IQVIA DA GERMANY: IQVIA Disease Analyser Germany, IQVIA LPD France: IQVIA Longitudinal Patient Data France, IPCI: Integrated Primary Care Information, IQVIA LPD Italy: IQVIA Longitudinal Patient Data Italy, SIDIAP CMBDH-HA: Information System for Research in Primary Care with hospital linkage. COPD: chronic obstructive pulmonary disease

Incidence rates for the different outcomes of thrombosis and TTS across the databases are summarised in Table 2. The incidence rates for CVST ranged from 0.3 (95% CI: 0.2 to 0.5) to 1.2 (1.0 to 1.5) per 100,000 person-years; CVST with thrombocytopenia was only seen in SIDIAP CMBDH-HA, where the incidence rate was 0.1 (0.1 to 0.2) per 100,000 person-years. The incidence rates for SVT ranged from 1.5 (1.2 to 1.8) to 10.3 (9.8 to 10.8), and from 0.1 (0.0 to 0.1) to 2.5 (2.2 to 2.7) per 100,000 person-years for SVT with thrombocytopenia. The incidence rates for DVT ranged from 85.9 (84.6 to 87.2) to 187.2 (182.6 to 191.8), and from 1.0 (0.7 to 1.4) to 8.5 (7.4 to 9.9) per 100,000 person-years for DVT with thrombocytopenia. The incidence rates for PE ranged from 66.1 (63.0 to 69.2) to 131.2 (126.4to 136.2), and from 0.5 (0.3 to 0.6) to 20.8 (18.9 to 22.8) per 100,000 person-years for PE with thrombocytopenia. Lastly, the incidence rates for stroke ranged from 128.3 (126.0 to 130.6) to 326.6 (320.6 to 332.7), and from 0.6 (0.4 to 0.7) to 30.9 (28.6 to 33.3) per 100,000 person-years for stroke with thrombocytopenia. As with thrombosis in general, incidence rates for TTS were typically higher for older age groups, see Figure 1.

**Table 2.** Incidence rates per 100,000 person-years for thrombosis and thrombosis with thrombocytopenia

|  | N | PYs | Number of events | Incidence rate (95% CI) per 100,000 PYs |
| --- | --- | --- | --- | --- |
| *Cerebral venous sinus thrombosis (CVST)* |  |  |  |  |
| CPRD | 3,913,025 | 9,676,085 | 118 | 1.2 (1.0 to 1.5) |
| IQVIA DA Germany | 8,459,044 | 19,369,671 | 95 | 0.5 (0.4 to 0.6) |
| IQVIA LPD France | 3,951,606 | 8,210,128 | 26 | 0.3 (0.2 to 0.5) |
| SIDIAP CMBDH-HA | 5,794,764 | 16,751,651 | 121 | 0.7 (0.6 to 0.9) |
| *Cerebral venous sinus thrombosis (CVST) with thrombocytopenia* |  |  |  |  |
| SIDIAP CMBDH-HA | 5,794,777 | 16,751,791 | 16 | 0.1 (0.1 to 0.2) |
| *Deep vein thrombosis (DVT)* |  |  |  |  |
| CPRD | 3,909,649 | 9,656,721 | 9,071 | 93.9 (92.0 to 95.9) |
| HIC Dundee | 948,184 | 2,153,442 | 1,186 | 55.1 (52.0 to 58.3) |
| IQVIA DA Germany | 8,451,032 | 19,329,175 | 16,600 | 85.9 (84.6 to 87.2) |
| IPCI | 1,296,310 | 3,402,027 | 6,367 | 187.2 (182.6 to 191.8) |
| IQVIA LPD Italy | 1,063,587 | 2,639,975 | 3,891 | 147.4 (142.8 to 152.1) |
| SIDIAP CMBDH-HA | 5,790,802 | 16,722,842 | 14408 | 86.2 (84.8 to 87.6) |
| *Deep vein thrombosis (DVT) with thrombocytopenia* |  |  |  |  |
| CPRD | 3,913,031 | 9,676,132 | 127 | 1.3 (1.1 to 1.6) |
| HIC Dundee | 948,498 | 2,153,995 | 184 | 8.5 (7.4 to 9.9) |
| IQVIA DA Germany | 8,458,995 | 19,369,390 | 225 | 1.2 (1.0 to 1.3) |
| IPCI | 1,299,274 | 3,418,833 | 34 | 1.0 (0.7 to 1.4) |
| IQVIA LPD Italy | 1,066,209 | 2,651,714 | 39 | 1.5 (1.0 to 2.0) |
| SIDIAP CMBDH-HA | 5,794,559 | 16,750,224 | 1037 | 6.2 (5.8 to 6.6) |
| *Pulmonary embolism (PE)* |  |  |  |  |
| CPRD | 3,910,531 | 9,662,585 | 7149 | 74.0 (72.3 to 75.7) |
| HIC Dundee | 947,984 | 2,151,351 | 2823 | 131.2 (126.4 to 136.2) |
| IQVIA DA Germany | 8,449,246 | 19,325,600 | 17204 | 89.0 (87.7 to 90.4) |
| IQVIA LPD France | 3,947,450 | 8,195,137 | 4700 | 57.4 (55.7 to 59.0) |
| IPCI | 1,297,807 | 3,410,984 | 3141 | 92.1 (88.9 to 95.4) |
| IQVIA LPD Italy | 1,064,532 | 2,645,601 | 1748 | 66.1 (63.0 to 69.2) |
| SIDIAP CMBDH-HA | 5,792,195 | 16,734,624 | 9590 | 57.3 (56.2 to 58.5) |
| *Pulmonary embolism (PE) with thrombocytopenia* |  |  |  |  |
| CPRD | 3,913,042 | 9,676,256 | 84 | 0.9 (0.7 to 1.1) |
| HIC Dundee | 948,459 | 2,153,685 | 447 | 20.8 (18.9 to 22.8) |
| DA Germany | 8,458,971 | 19,369,265 | 286 | 1.5 (1.3 to 1.7) |
| IQVIA LPD France | 3,951,605 | 8,210,109 | 39 | 0.5 (0.3 to 0.6) |
| IPCI | 1,299,282 | 3,418,860 | 21 | 0.6 (0.4 to 0.9) |
| IQVIA LPD Italy | 1,066,222 | 2,651,761 | 17 | 0.6 (0.4 to 1.0) |
| SIDIAP CMBDH-HA | 5,794,594 | 16,750,449 | 985 | 5.9 (5.5 to 6.3) |
| *Splanchnic vein thrombosis (SVT)* |  |  |  |  |
| CPRD | 3,913,005 | 9,675,960 | 233 | 2.4 (2.1 to 2.7) |
| HIC Dundee | 948,541 | 2,154,020 | 112 | 5.2 (4.3 to 6.3) |
| IQVIA DA Germany | 8,458,941 | 19,369,177 | 398 | 2.1 (1.9 to 2.3) |
| IQVIA LPD France | 3,951,594 | 8,210,016 | 122 | 1.5 (1.2 to 1.8) |
| IQVIA LPD Italy | 1,066,207 | 2,651,684 | 58 | 2.2 (1.7 to 2.8) |
| SIDIAP CMBDH-HA | 5,794,483 | 16,749,703 | 1718 | 10.3 (9.8 to 10.8) |
| *Splanchnic vein thrombosis (SVT) with thrombocytopenia* |  |  |  |  |
| CPRD | 3,913,070 | 9,676,375 | 5 | 0.1 (0.0 to 0.1) |
| HIC Dundee | 948,553 | 2,154,098 | 40 | 1.9 (1.3 to 2.5) |
| IQVIA DA Germany | 8,459,086 | 19,369,887 | 16 | 0.1 (0.0 to 0.1) |
| SIDIAP CMBDH-HA | 5,794,721 | 16,751,354 | 412 | 2.5 (2.2 to 2.7) |
| *Stroke* |  |  |  |  |
| CPRD | 3,908,602 | 9,650,786 | 12381 | 128.3 (126.0 to 130.6) |
| HIC Dundee | 947,200 | 2,146,633 | 6,326 | 294.7 (287.5 to 302.0) |
| IQVIA DA Germany | 8,434,716 | 19,257,348 | 46784 | 242.9 (240.7 to 245.2) |
| IQVIA LPD France | 3,944,166 | 8,178,421 | 12899 | 157.7 (155.0 to 160.5) |
| IPCI | 1,293,518 | 3,390,322 | 11073 | 326.6 (320.6 to 332.7) |
| SIDIAP CMBDH-HA | 5,782,776 | 16,671,995 | 42126 | 252.7 (250.3 to 255.1) |
| *Stroke with thrombocytopenia* |  |  |  |  |
| CPRD | 3,913,039 | 9,676,223 | 79 | 0.8 (0.6 to 1.0) |
| HIC Dundee | 948,431 | 2,153,504 | 665 | 30.9 (28.6 to 33.3) |
| IQVIA DA Germany | 8,458,923 | 19,369,062 | 369 | 1.9 (1.7 to 2.1) |
| IQVIA LPD France | 3,951,616 | 8,210,137 | 46 | 0.6 (0.4 to 0.7) |
| IPCI | 1,299,278 | 3,418,845 | 32 | 0.9 (0.6 to 1.3) |
| SIDIAP CMBDH-HA | 5,794,204 | 16,747,541 | 3058 | 18.3 (17.6 to 18.9) |

CPRD: Clinical Practice Research Datalink, IQVIA DA GERMANY: IQVIA Disease Analyser Germany, IQVIA LPD France: IQVIA Longitudinal Patient Data France, IPCI: Integrated Primary Care Information, IQVIA LPD Italy: IQVIA Longitudinal Patient Data Italy, SIDIAP CMBDH-HA: Information System for Research in Primary Care with hospital linkage.

**Figure 1.**
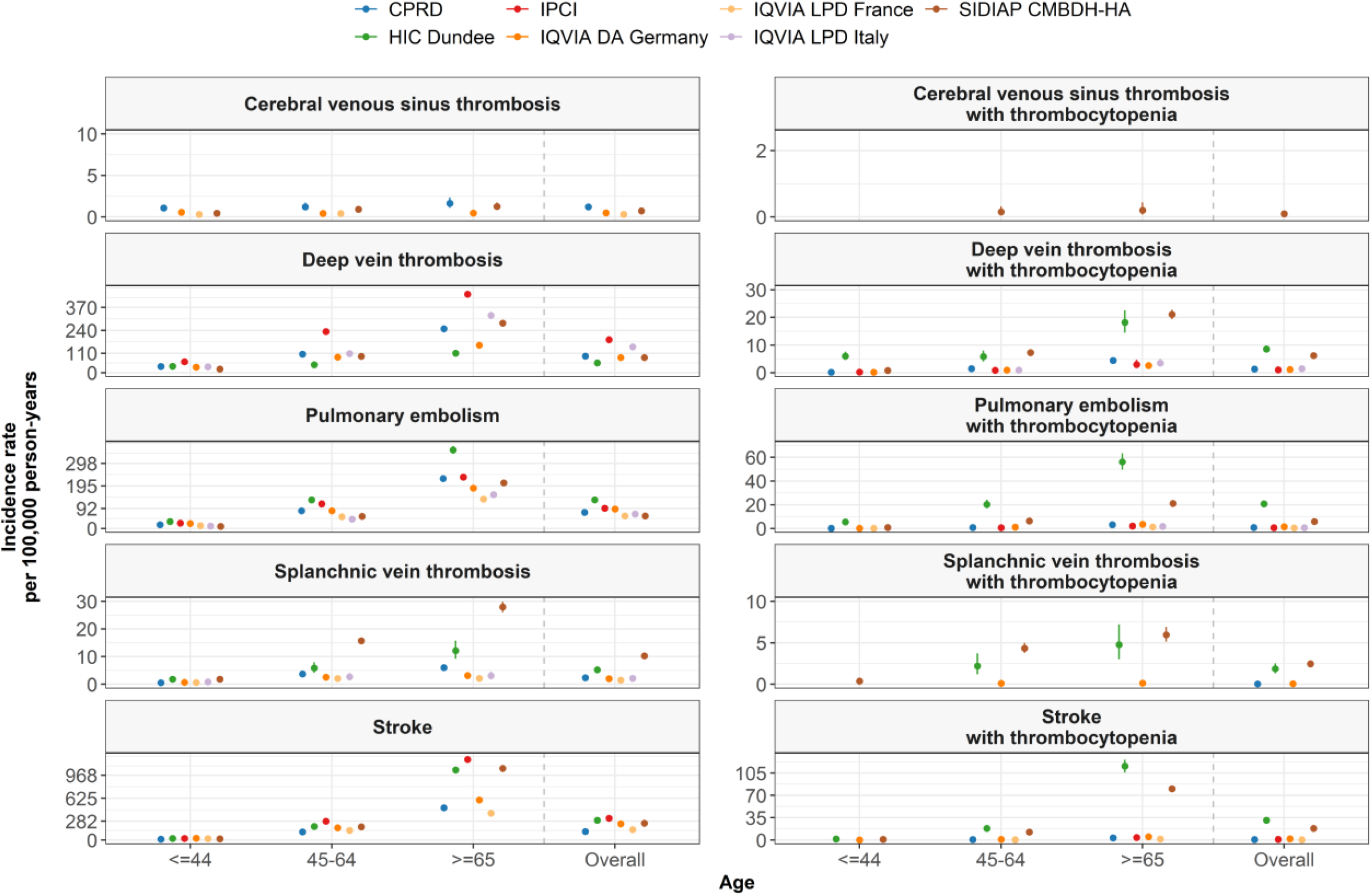
Incidence rates (with 95% confidence intervals) per 100,000 of TTS among the general population, stratified by age and sex

Based on the highest estimates for the overall study cohorts, one would expect approximately 1 case of CVST with thrombocytopenia, 19 of SVT with thrombocytopenia, 65 of DVT with thrombocytopenia, 159 of PE with thrombocytopenia, and 237 of stroke with thrombocytopenia among a general population of 10 million individuals per 28 days. For a cohort of the same size aged 65 or over, this would rise to 46 of SVT with thrombocytopenia, 161 of DVT with thrombocytopenia, 886 of PE with thrombocytopenia, and 616 of stroke with thrombocytopenia, see Figure 2.

**Figure 2.**
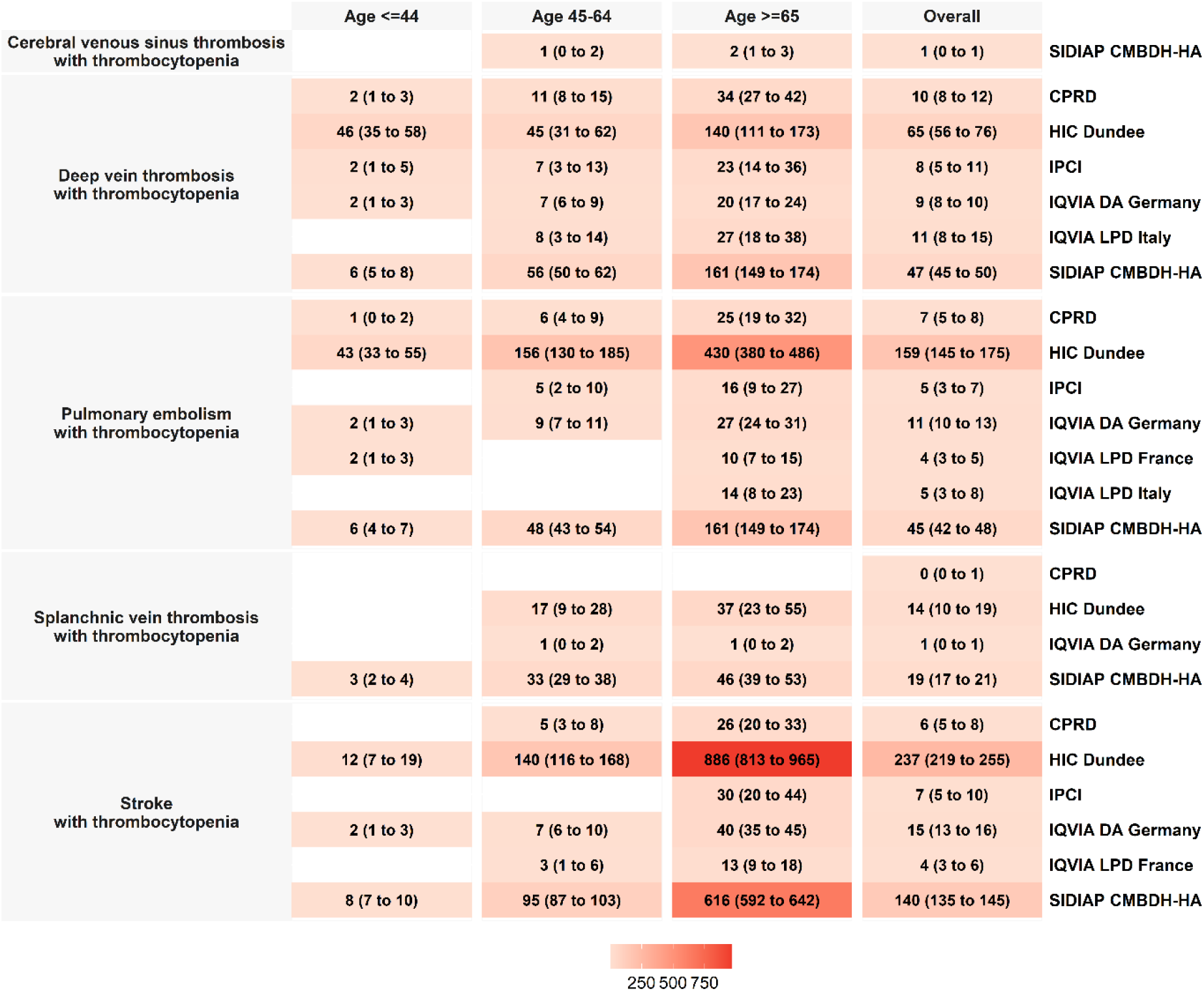
Expected cases (with 95% confidence intervals) of TTS per 28 days in a population of 10,000,000 people in a given age strata or overall. Blank cells are where there were fewer than five people with the event and incidence rates were not estimated.

The age and sex profiles of those with TTS are summarised in Table 3 and the prevalence of comorbidities and prior medication are presented in Figure 3, along with those of the study populations. The median age of the 16 individuals with CVST with thrombocytopenia in SIDIAP CMBDH-HA was 62 years old. The median age of those with DVT with thrombocytopenia ranged from 58 to 76 across the databases, from 68 to 78 for PE with thrombocytopenia, from 59 to 64 for SVT with thrombocytopenia, and from 73 to 78 for stroke with thrombocytopenia. Men generally predominated the affected cohorts, accounting for 50.0% to 71.6% of those with different TTS in the contributing databases. The prevalence of comorbidities and prior medication use was higher for patients with TTS than in the general population. In CPRD, for example, 1.8% of the study population had an autoimmune disease, 5.1% had a history of cancer, 5.5% had diabetes, 4.3% had renal impairment. These compared to 12.6%, 25.2%, 20.5%, and 26.8% for patients with DVT with thrombocytopenia. Similarly, while 2.9% of the study population were taking antithrombotic and anticoagulant therapies in the months preceding their index date, 18.1% of patients with DVT with thrombocytopenia were. Requiring a year of prior history for study participants to be included in the analysis and defining study populations based on their first visit after 2017 had only a small effect on the results, see the Appendix.

**Table 3.** Characteristics of patients with thrombosis with thrombocytopenia

|  | N | Age (Median [IQR]) | Sex: Male (N [%]) |
| --- | --- | --- | --- |
| *CPRD* |  |  |  |
| Study population | 3,913,071 | 41 [22 to 59] | 1,937,858 (49.5%) |
| Deep vein thrombosis with thrombocytopenia | 127 | 70 [56 to 80] | 70 (55.1%) |
| Pulmonary embolism with thrombocytopenia | 84 | 71 [62 to 79] | 50 (59.5%) |
| Splanchnic vein thrombosis with thrombocytopenia | 5 | 59 [52 to 59] | <5 |
| Stroke with thrombocytopenia | 79 | 74 [66 to 81] | 53 (67.1%) |
| *HIC Dundee* |  |  |  |
| Study population | 948,561 | 41 [23 to 59] | 469,725 (49.5%) |
| Deep vein thrombosis with thrombocytopenia | 184 | 58 [37 to 75] | 99 (53.8%) |
| Pulmonary embolism with thrombocytopenia | 447 | 68 [53 to 78] | 247 (55.3%) |
| Splanchnic vein thrombosis with thrombocytopenia | 40 | 65 [52 to 72] | 27 (67.5%) |
| Stroke with thrombocytopenia | 665 | 77 [67 to 84] | 398 (59.8%) |
| *IQVIA DA Germany* |  |  |  |
| Study population | 8,459,098 | 52 [32 to 67] | 3,589,506 (42.4%) |
| Deep vein thrombosis with thrombocytopenia | 225 | 71 [60 to 80] | 143 (63.6%) |
| Pulmonary embolism with thrombocytopenia | 286 | 72 [62 to 80] | 183 (64.0%) |
| Splanchnic vein thrombosis with thrombocytopenia | 16 | 64 [60 to 73] | 11 (68.8%) |
| Stroke with thrombocytopenia | 369 | 76 [66 to 82] | 244 (66.1%) |
| *IQVIA LPD France* |  |  |  |
| Study population | 3,951,633 | 48 [28 to 65] | 1,669,415 (42.2%) |
| Pulmonary embolism with thrombocytopenia | 39 | 72 [57 to 83] | 23 (59.0%) |
| Stroke with thrombocytopenia | 46 | 73 [66 to 82] | 30 (65.2%) |
| *IPCI* |  |  |  |
| Study population | 1,299,288 | 44 [23 to 60] | 636,386 (49.0%) |
| Deep vein thrombosis with thrombocytopenia | 34 | 70 [54 to 81] | 20 (58.8%) |
| Pulmonary embolism with thrombocytopenia | 21 | 70 [54 to 73] | 12 (57.1%) |
| Stroke with thrombocytopenia | 32 | 74 [68 to 85] | 22 (68.8%) |
| *IQVIA LPD Italy* |  |  |  |
| Study population | 1,066,230 | 52 [37 to 68] | 426,758 (40.0%) |
| Deep vein thrombosis with thrombocytopenia | 39 | 76 [62 to 82] | 20 (51.3%) |
| Pulmonary embolism with thrombocytopenia | 17 | 78 [69 to 81] | 9 (52.9%) |
| *SIDIAP CMBDH-HA* |  |  |  |
| Study population | 5,794,777 | 42 [25 to 59] | 2,859,044 (49.3%) |
| Cerebral venous sinus thrombosis with thrombocytopenia | 16 | 62 [49 to 67] | 8 (50.0%) |
| Deep vein thrombosis with thrombocytopenia | 1,037 | 69 [58 to 79] | 617 (59.5%) |
| Pulmonary embolism with thrombocytopenia | 985 | 70 [59 to 79] | 584 (59.3%) |
| Splanchnic vein thrombosis with thrombocytopenia | 412 | 62 [54 to 72] | 295 (71.6%) |
| Stroke with thrombocytopenia | 3,058 | 76 [67 to 83] | 2,005 (65.6%) |

CPRD: Clinical Practice Research Datalink, IQVIA DA GERMANY: IQVIA Disease Analyser Germany, IQVIA LPD France: IQVIA Longitudinal Patient Data France, IPCI: Integrated Primary Care Information, IQVIA LPD Italy: IQVIA Longitudinal Patient Data Italy, SIDIAP CMBDH-HA: Information System for Research in Primary Care with hospital linkage

**Figure 3.**
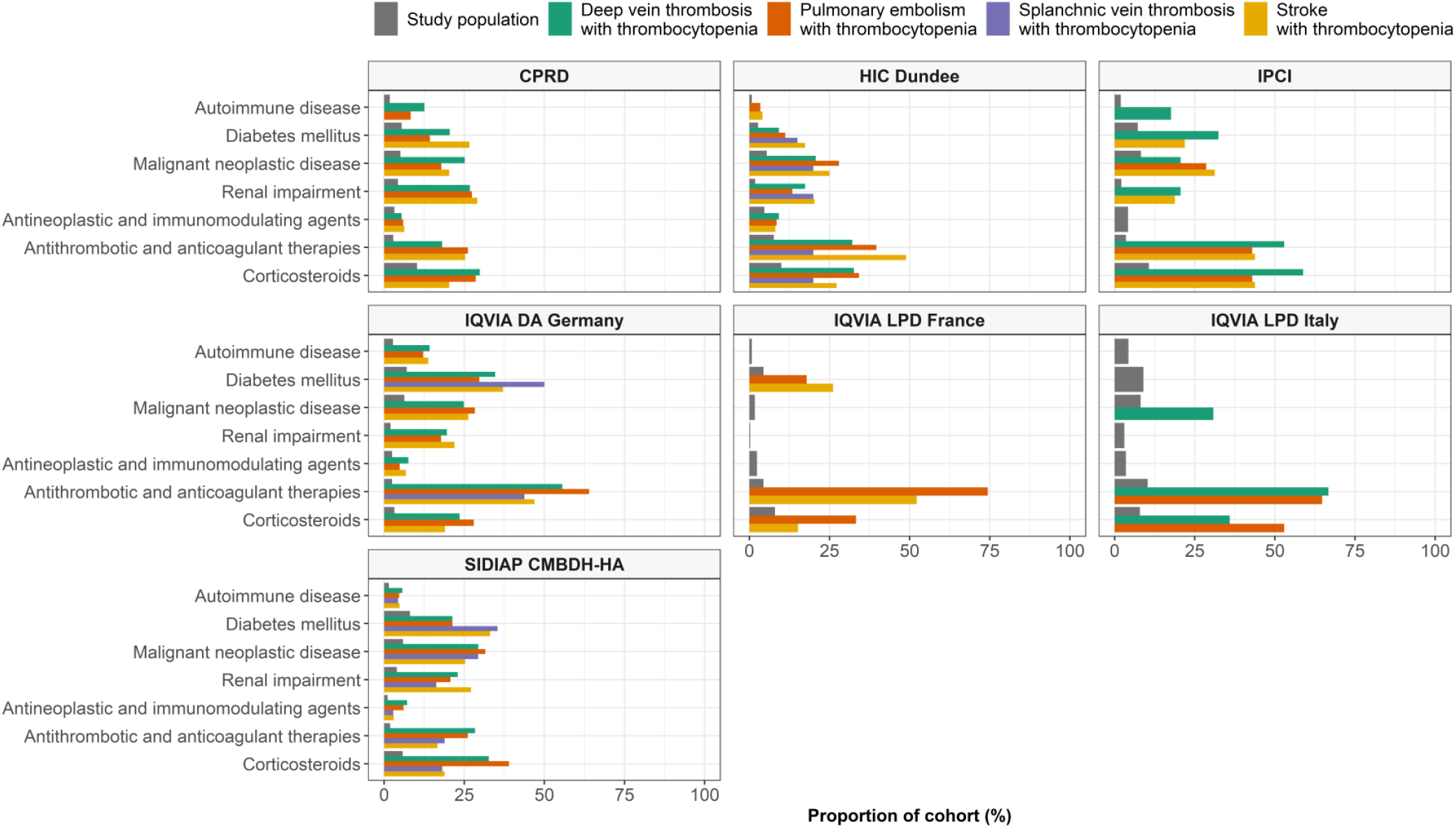
Comorbidities and prior medication use among patients with TTS compared to the overall study population. Any characteristic seen in less than five people in a cohort is not reported.

Incidence rates for DIC, HIT, immune thrombocytopenia, and TTP are summarised in Table 4. The incidence rate for DIC ranged from 0.2 (0.1 to 0.3) to 3.8 (3.3 to 4.1) per 100,000 person-years, from 0.2 (0.1 to 0.4) to 37.9 (37.0 to 38.) for HIT, from 2.1 (1.8 to 2.5) to 46.7 (45.7 to 47.7) for immune thrombocytopenia, and from 0.4 (0.2 to 0.8) to 2.8 (2.6 to 3.1) for thrombotic thrombocytopenic purpura.

**Table 4.** Incidence rates per 100,000 person-years for coagulopathy

|  | N | PYs | Number of events | Incidence rate per 100, 000 PYs |
| --- | --- | --- | --- | --- |
| *Disseminated intravascular coagulation* |  |  |  |  |
| CPRD | 3,913,067 | 9,676,361 | 15 | 0.2 (0.1 to 0.3) |
| HIC Dundee | 948,555 | 2,154,105 | 32 | 1.5 (1.0 to 2.1) |
| IQVIA DA Germany | 8,459,041 | 19,369,706 | 79 | 0.4 (0.3 to 0.5) |
| IQVIA France LPD | 3,951,611 | 8,210,148 | 34 | 0.4 (0.3 to 0.6) |
| IQVIA Italy LPD | 1,066,206 | 2,651,697 | 37 | 1.4 (1.0 to 1.9) |
| SIDIAP CMBDH-HA | 5,794,596 | 16,750,823 | 641 | 3.8 (3.5 to 4.1) |
| *Heparin-induced thrombocytopenia* |  |  |  |  |
| CPRD | 3,912,943 | 9,675,822 | 302 | 3.1 (2.8 to 3.5) |
| HIC Dundee | 948,500 | 2,153,919 | 165 | 7.7 (6.5 to 8.9) |
| IQVIA DA Germany | 8,458,456 | 19,366,573 | 1513 | 7.8 (7.4 to 8.2) |
| IQVIA France LPD | 3,951,623 | 8,210,192 | 20 | 0.2 (0.1 to 0.4) |
| IQVIA Italy LPD | 1,066,144 | 2,651,425 | 171 | 6.4 (5.5 to 7.5) |
| SIDIAP CMBDH-HA | 5,792,945 | 16,739,615 | 6351 | 37.9 (37.0 to 38.9) |
| *Immune thrombocytopenia* |  |  |  |  |
| CPRD | 3,912,708 | 9,674,616 | 759 | 7.8 (7.3 to 8.4) |
| HIC Dundee | 948,447 | 2,153,668 | 333 | 15.5 (13.8 to 17.2) |
| IQVIA DA Germany | 8,457,949 | 19,364,321 | 2264 | 11.7 (11.2 to 12.2) |
| IQVIA LPD France | 3,951,527 | 8,209,807 | 175 | 2.1 (1.8 to 2.5) |
| IPCI | 1,299,133 | 3,418,075 | 267 | 7.8 (6.9 to 8.8) |
| SIDIAP CMBDH-HA | 5,792,354 | 16,736,041 | 7816 | 46.7 (45.7 to 47.7) |
| *Thrombotic thrombocytopenic purpura* |  |  |  |  |
| CPRD | 3,913,059 | 9,676,289 | 52 | 0.5 (0.4 to 0.7) |
| HIC Dundee | 948,560 | 2,154,123 | 9 | 0.4 (0.2 to 0.8) |
| IQVIA DA Germany | 8,458,805 | 19,368,465 | 552 | 2.8 (2.6 to 3.1) |
| IQVIA LPD France | 3,951,599 | 8,210,086 | 52 | 0.6 (0.5 to 0.8) |
| IQVIA LPD Italy | 1,066,176 | 2,651,585 | 45 | 1.7 (1.2 to 2.3) |
| SIDIAP CMBDH-HA | 5,794,674 | 16,751,190 | 272 | 1.6 (1.4 to 1.8) |

CPRD: Clinical Practice Research Datalink, IQVIA DA GERMANY: IQVIA Disease Analyser Germany, IQVIA LPD France: IQVIA Longitudinal Patient Data France, IPCI: Integrated Primary Care Information, IQVIA LPD Italy: IQVIA Longitudinal Patient Data Italy, SIDIAP CMBDH-HA: Information System for Research in Primary Care with hospital linkage

The incidence rates for all study outcomes are summarised in the supplementary materials and in a web application: https://livedataoxford.shinyapps.io/CovCoagBackgroundIncidence/, where the characteristics of outcome cohorts are also described.

## Discussion

### Key results

In this study we have analysed data for over 25 million people from across 6 European countries to establish the background incidence of TTS. With incidence rates of less than 35 per 100,000 person-years, TTS can be considered as a very rare event. These events can generally be expected to occur in older persons, with the average age of those over 60 for most events in most of the databases studied. Moreover, those affected typically had a higher prevalence of comorbidities, such as autoimmune diseases, cancer, and diabetes. They also had a high prevalence of use of medications indicated for the prevention of thrombosis including antithrombotic and anticoagulant therapies, as well as some potentially associated with an increased risk of TTS such as systemic glucocorticoids. Coagulopathies potentially associated with TTS were mostly rare: immune thrombocytopenia was the most common with rates up to almost 47 per 100,000 person-years, followed by HIT (up to 38 per 100,000), DIC (up to 4 per 100,000), and TTP (up to 3 per 100,000).

Exact estimates of the background incidence of both rare events, such as CVST, and more common events, such as DVT, have though been seen to vary substantially depending on the data source used. This comes even after the use of common analytic code and with databases mapped to a common data model. More specifically, the databases which captured both inpatient and outpatient records showed the highest background rates of most of the studied events. Moreover, rates of TTS were much higher in the database which captured both inpatient and outpatient lab measurements (HIC Dundee). Indeed, while around 90% of those with VTE had a platelet count recorded in this database, only around 10% had a count in the other databases which relied on outpatient lab measurements. In both cases, around 10% of lab measurements that were observed had a value that indicated thrombocytopenia. This underlines the importance of using consistent data sources in vaccine safety research.

### Findings in context

A number of previous studies have estimated the incidence of venous thromboembolism in the general population, with its incidence rate estimated to be around 100 persons per 100,000 person-years.^18^ Approximately two-thirds of venous thromboembolism can be expected to present as DVT, with the other third presenting as PE with or without DVT.^19^ The background of stroke has also been the subject of much research, with its incidence generally estimated to be more than 100 persons per 100,000 person-years. ^20,21^ The incidence of each of these events is seen to be much higher among older persons. The incidence of SVT and CVST is far less well-known. Estimates for the incidence of CVST have ranged from 0.2 to 2 per 100,000 person-years. ^22–25^ Meanwhile there is little research describing the incidence of SVT in the general population, although the incidence of portal vein thrombosis, the most commonly involved vein, has been estimated at around 3 per 100,000 person-years, while the incidence of Budd-Chiari syndrome was estimated at around 2 per 100,000 person-years in the same study.^26^

In a recent study data from Denmark and Norway was used to assess 28-day rates of thromboembolic events and coagulation disorders among a cohort of people who had received the ChAdOx1 vaccine and in historical comparator cohorts.^27^ In the historical comparator population, which covered 2016 to 2018 for Denmark and 2018 to 2019 for Denmark, the incidence rate of CVST, PE, lower limb venous thrombosis, and SVT were estimated at 2, 57, 94, and 4 per 100,000 person-years, respectively. Meanwhile the incidence rate for idiopathic thrombocytopenia purpura and DIC were 7 and 1 per 100,000. These estimates are broadly in line with those from our study.

The background incidence of TTS itself, however, has not been previously described in detail. Our findings demonstrate that thrombosis with concomitant thrombocytopenia is very rare, with an unadjusted overall incidence rate of less than 5 per 100,000 person-years for each of DVT, PE, CVST, SVT, and stroke with thrombocytopenia. In particular, CVST with thrombocytopenia was only seen in the database with patient-level linkage to hospital data, where its incidence rate was approximately one per million person-years. Similarly, the incidence rate for SVT with thrombocytopenia was between one and seven cases per million.

Spontaneous identified 93 cases of CVST with thrombocytopenia among individuals who had recently received the ChAdOx1 vaccine in the UK.^8^ The profile of patients with TTS after vaccination also appears to differ to the typical profiles of those with TTS as seen in our data. While in this study we have seen those with TTS to typically be older than the general population, more commonly male, and with more comorbidities and greater prior medication use, initial studies describing the profiles of patients with vaccine-induced TTS have most often presented the cases of people who were aged under 60, more often female, and with relatively few comorbidities described.^28–31^ While these case series of TTS after vaccination are small and their profiles may reflect the particular characteristics of those who were first to receive a vaccine against SARS-CoV-2, this dissimilarity in patient profiles of those with TTS in previous years and those for whom it has been reported following a vaccination is notable.

Where we have estimated the incidence of thrombosis in general, our findings have been very broadly in line with previous research. Estimates have, however, been seen to vary substantially across databases. This was particularly the case for estimates for DVT, for example, where a three-fold difference was seen between the databases with the highest and lowest incidence rates. Our estimates of TTS and coagulopathy also vary across the databases used. Indeed, this heterogeneity was observed although we used data mapped to a common data model and applied the same analytic code across the databases. Given that the data sources used come from different countries and can be expected to vary in completeness, accuracy, and granularity, it is not necessarily surprising that estimates vary. In addition, given the seriousness of many of the events described here, the degree to which episodes of hospital care are reflected in the data is also likely to be an important driver of the observed heterogeneity, with incidence rates higher for most of the events in the data source with linkage to hospitalisation data. The reasons for the heterogeneity seen between the databases used in this study is discussed in further detail in the Appendix.

### Study limitations

This study relies on routinely-collected health care data and while this has allowed for the inclusion of a large study population, the recording of TTS has not previously been evaluated in the databases used. A degree of measurement error can thus be expected, and further research is required to validate the recording of TTS.

The degree to which the TTS events being described after vaccinations against SARS-CoV-2 are comparable to TTS events previously seen in the general population is as yet unclear. TTS after vaccination appears to occur at unusual sites, with a large proportion of spontaneous reports and case series describing cerebral or abdominal thromboses, and with high levels of antibodies to platelet factor 4 often observed despite the absence of an exposure to heparin.^28,32^ In this study we have focused on specific sites of thrombosis with concomitant thrombocytopenia. We believe that this is more instructive than providing a singular background incidence rate for venous thromboembolism with thrombocytopenia, which would be driven in large part by commonly seen events (such as DVT and PE) and would not necessarily reflect the presentation of TTS after vaccination. As the pathophysiology of TTS after vaccination becomes better understood, definitions of the appropriate historical comparator can also be expected to evolve so as to best match the condition being described among those who have been recently vaccinated.

### Conclusion

Based on data from over 25 million people from six European databases, TTS has been seen to be very rare. While rates varied across databases, the highest incidence rates for DVT, PE, and stroke with thrombocytopenia were 8.5, 20.8, and 30.9 per 100,000 person-years, respectively. Meanwhile the highest incidence rates for CVST and SVT with thrombocytopenia were 0.1 and 2.5 per 100,000 person-years. TTS was typically seen among individuals older, more often male, and in worse health than the general population. While these findings help to provide context for the rates of adverse events being reported by spontaneous reports following vaccinations against SARS-CoV-2, a full assessment of the safety signal for TTS would benefit from within-database comparisons which account for individual-level characteristics such as age and sex.

## Ethical approvals

The protocol for this research was approved by the Independent Scientific Advisory Committee (ISAC) for MHRA Database Research (protocol number 20_000211), the IDIAPJGol Clinical Research Ethics Committee (project code: 21/007-PCV), and the IPCI governance board (application number 3/2021). Some databases used (IQVIA LPD Italy, IQVIA LPD France, IQVIA DA Germany) in these analyses are commercially available, syndicated data assets that are licensed by contributing authors for observational research. These assets are de-identified commercially available data products that could be purchased and licensed by any researcher. As these data are deemed commercial assets, there is no Institutional Review Board applicable to the usage and dissemination of these result sets or required registration of the protocol with additional ethics oversight. Compliance with Data Use Agreement terms, which stipulate how these data can be used and for what purpose, is sufficient for the licensing commercial entities. Further inquiry related to the governance oversight of these assets can be made with the respective commercial entity, IQVIA (iqvia.com). For HIC Dundee, institutional review board approval for the use of de-identified data for this project was granted by the Tayside Health Informatics Centre.

## Acknowledgements

This study was funded by the European Medicines Agency in the form of a competitive tender (Lot ROC No EMA/2017/09/PE). We acknowledge Prof Johan Van der Lei for the overall management of this research grant.

## Author contributions

All authors were involved in the study conception and design, interpretation of the results, and the preparation of the manuscript. EB led the data analysis and wrote the initial draft of the manuscript with DPA. EB, TDS, CR, MA, and SFB had access to the SIDIAP data, EB, XL, AD, and DPA had access to the CPRD data, DM and SH had access to the HIC Dundee data, PR had access to the IPCI data, and KK, HMS, CR, SS, had access to LPD France, LPD Italy, and DA Germany.

## Appendix

### A1. Impact of requiring a year of prior history for study participants to be included in the analysis

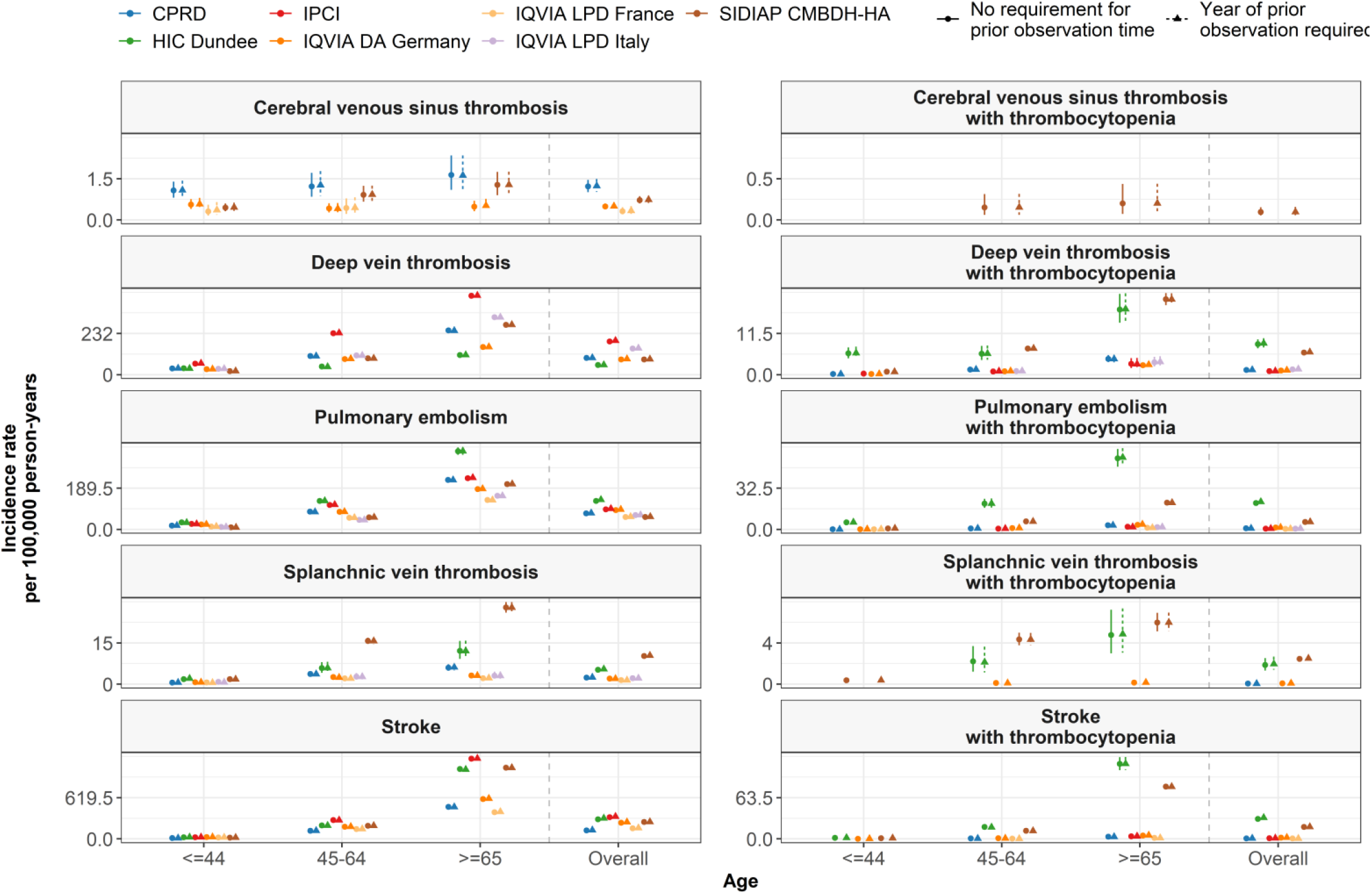

### A2. Impact of defining study populations based on a visit

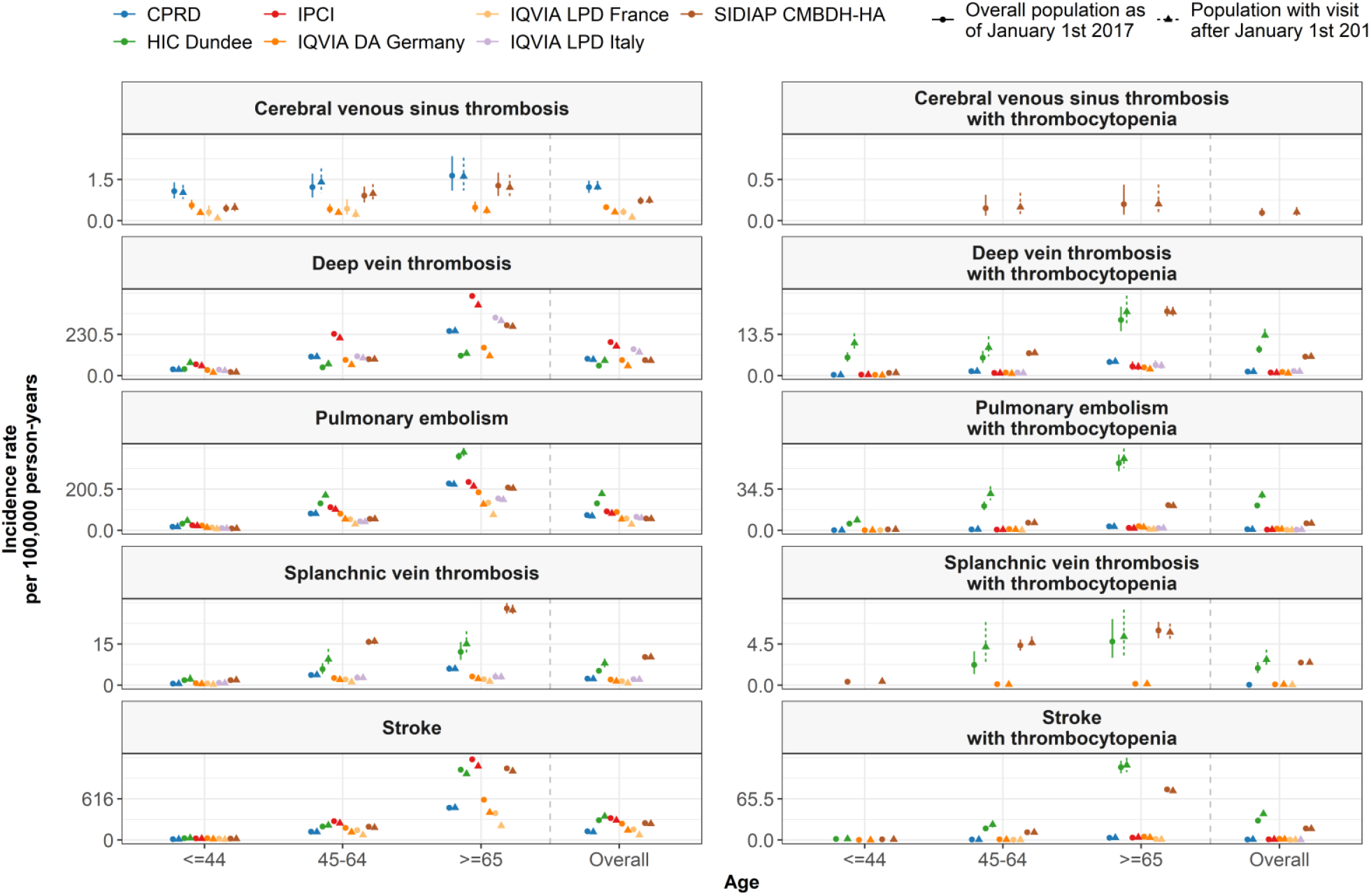

### A3. Heterogeneity in the incidence rates across databases

In this study variation in incidence rates has been seen across the databases used. Below we discuss the reasons for this, with a particular focus on the results for deep vein thrombosis as an example.

#### Analytic choices

While heterogeneity between studies in the literature can often in part be explained by variation in analytic choices, this is not the case in this study where we applied the same analytic code to each database.

#### Differences in setting

Variation in the ‘strue’ background incidence rates across countries is not in itself unexpected, given differences in population demographics, the prevalence of risk factors for the events of interest, and so on. For example, risk factors for deep vein thrombosis include obesity (https://www.sciencedirect.com/science/article/pii/S0140673605718808), and the prevalence of obesity is well-known to vary substantially across countries (https://link.springer.com/article/10.1007/s00394-014-0746-4).

#### Differences in source coding systems

Medical vocabularies vary and the databases used in the study use Read codes (CPRD), ICPC (IPCI), ICD-9 (LPD Italy), ICD-10 (DA Germany and LPDD France), and ICD-10CM (SIDIAP) to represent condition-related concepts. These coding systems differ in the way that they describe clinical events (in particular their granularity) and this can have a meaningful impact on research findings. For example, this can be seen in the literature by the impact on research findings when databases switched from using ICD-9 to ICD-10 codes. (https://onlinelibrary.wiley.com/doi/abs/10.1002/pds.4563).

In our study, a substantial impact can be seen for some databases when using the narrow definition for DVT rather than the broad one, see Figure 1 and Figure 2 below. The most common source codes are for our narrow DVT definition are “Deep vein thrombosis” (CPRD – Read code), “Phlebitis and thrombophlebitis of other deep vessels of lower extremities” (DA Germany and LPD France – ICD-10 code), “Other venous embolism and thrombosis of inferior vena cava” (LPD Italy-ICD-9 code), and “Phlebitis and thrombophlebitis of unspecified deep vessels of unspecified lower extremity” (SIDIAP – ICD-10-CM). While broadening the definition of DVT had only modest effect on the results for CPRD and IPCI, in did have a substantial impact for DA Germany (where the most common included code became “Phlebitis and thrombophlebitis of lower extremities, unspecified”), for LPD France (where the most common included code became “Phlebitis and thrombophlebitis of unspecified site”), for LPD Italy (where the most common included code became “Other venous embolism and thrombosis of unspecified site”), and SIDIAP (where the most common included code became “Phlebitis and thrombophlebitis of unspecified deep vessels of unspecified lower extremity”). This provides one example of the underlying variation between the databases in how the presentation of a particular is captured in the medical record. While using a common data model facilitates the analysis of the data sources in a standardised manner, this underlying heterogeneity in the source data is still reflected to an important degree.

**Figure A3.1.**
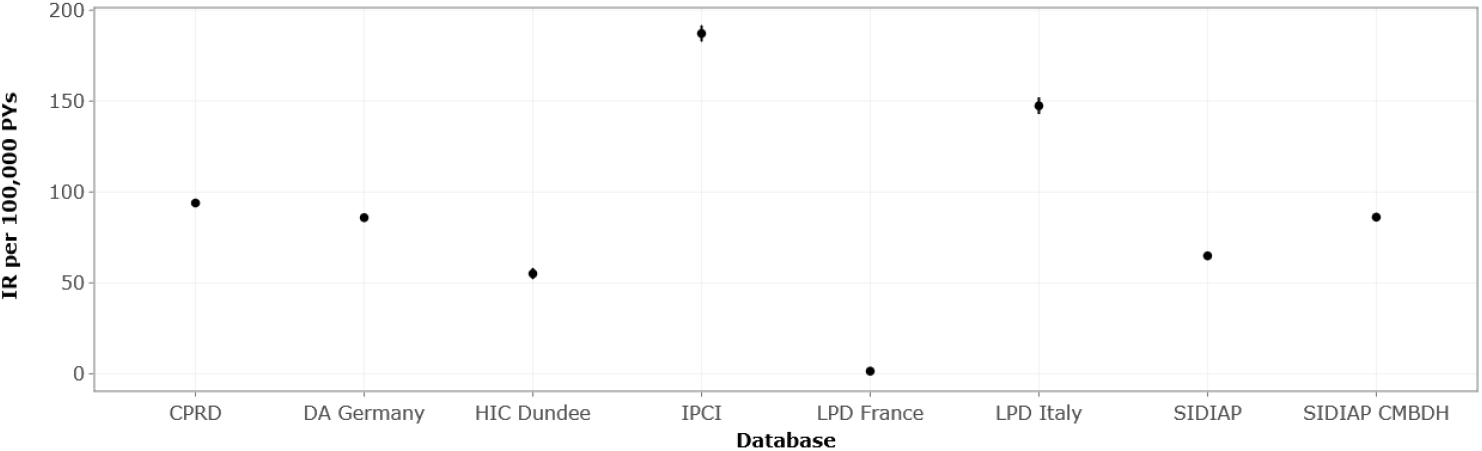
Estimated incidence rates for DVT using a narrow definition.

**Figure A3.2.**
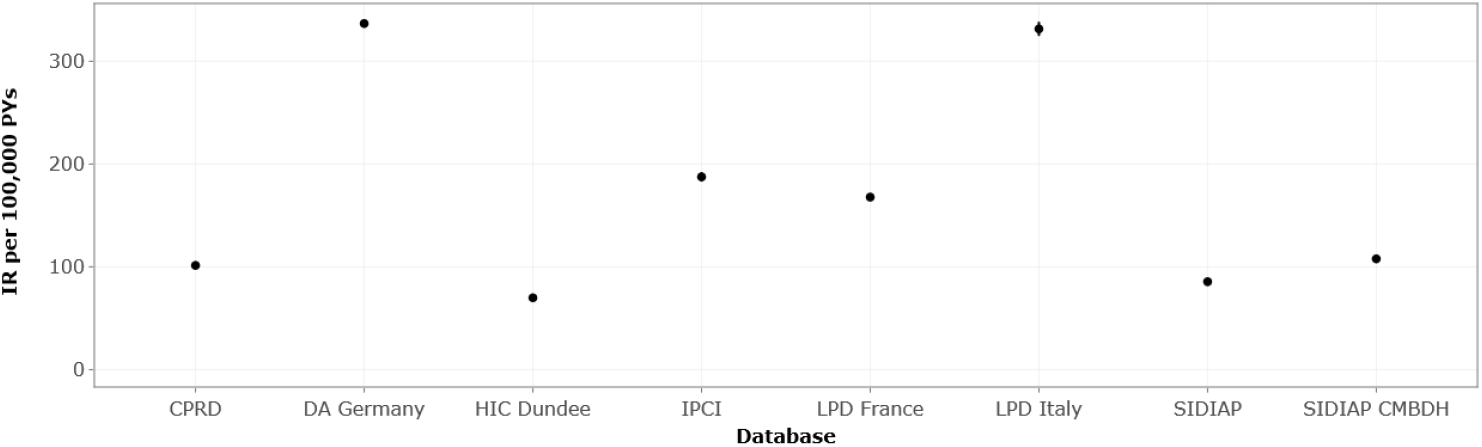
Estimated incidence rates for DVT using a broad definition.

#### Differences in capture of hospital events

Many of the events considered in our study can be expected to involve hospitalisation. Consequently, the degree to which diagnoses made in the hospital setting are reflected in the data sources used can be expected to have a large effect on the capture of events. We see this for SIDIAP where we ran the analysis in both the database with only primary care records and again with linkage at a patient-level to hospital records, SIDIAP CMBDH-HA.

An individual in the community with DVT may require hospital treatment and DVT may also occur in persons hospitalised for other reasons. For our narrow definition, we see that the incidence rate of DVT increases from 64.9 per 100,000 person-years in SIDIAP to 86.2 per 100,000 person-years in SIDIAP CMBDH-HA, see figure 3.

While we can expect patient-level linkage of the other data sources to hospital level data to impact their results, it will not necessarily be to the same degree as in SIDIAP. In Catalonia, general practitioners are generally able to view patient hospital records during a consultation in a separate program to that used to view primary care records, with only the latter is captured in the SIDIAP database. Consequently, for patient care there is not a need to copy across what is seen in the system with the hospital record into the system with the primary care record which is captured by SIDIAP. This is not the case in the UK, however, where there is a need to add the information from letters received by general practitioners from secondary care facilities into the electronic medical record. Consequently, the impact of hospital linkage is likely to be context-dependent, as well as varying depending on the event under consideration.

**Figure A3.3.**
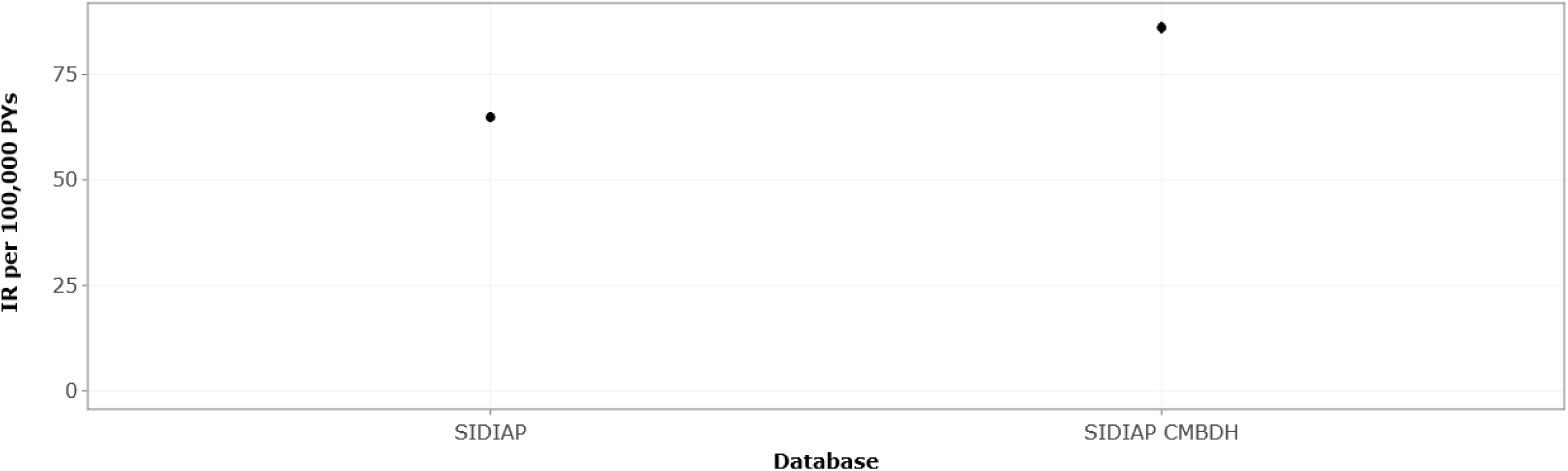
Estimated incidence rates for DVT using a narrow definition in SIDIAP and SIDIAP CMBDH-HA.

